# Predicting CoVID-19 community mortality risk using machine learning and development of an online prognostic tool

**DOI:** 10.1101/2020.04.27.20081794

**Authors:** Ashis Kumar Das, Shiba Mishra, Saji Saraswathy Gopalan

## Abstract

**Background:** The recent pandemic of CoVID-19 has emerged as a threat to global health security. There are a very few prognostic models on CoVID-19 using machine learning.

**Objectives:** To predict mortality among confirmed CoVID-19 patients in South Korea using machine learning and deploy the best performing algorithm as an open-source online prediction tool for decision-making.

**Materials and methods:** Mortality for confirmed CoVID-19 patients (n=3,299) between January 20, 2020 and April 30, 2020 was predicted using five machine learning algorithms (logistic regression, support vector machine, K nearest neighbor, random forest and gradient boosting). Performance of the algorithms was compared, and the best performing algorithm was deployed as an online prediction tool.

**Results:** The random forest algorithm was the best performer in terms of predictive ability (accuracy=0.981), discrimination (area under ROC curve=0.886), calibration (Matthews Correlation Coefficient=0.459; Brier Score=0.063) and. The best performer algorithm (random forest) was deployed as the online CoVID-19 Community Mortality Risk Prediction tool named CoCoMoRP (https://ashis-das.shinyapps.io/CoCoMoRP/).

**Conclusions:** We describe the development and deployment of an open-source machine learning tool to predict mortality risk among CoVID-19 confirmed patients using publicly available surveillance data. This tool can be utilized by potential stakeholders such as health providers and policy makers to triage patients at the community level in addition to other approaches.

## 1. Introduction

A novel coronavirus disease 2019 (CoVID-19) originated from Wuhan in China was reported to the World Health Organization in December of 2019.[1] Ever since, this novel coronavirus has spread to almost all major nations in the world resulting in a major pandemic. As of May 11, 2020, it has contributed to more than 4.1 million confirmed cases and about 283,000 deaths.[2] The first CoVID-19 case was diagnosed in South Korea on January 20, 2020. According to the Korea Centers for Disease Control and Prevention (KCDC), there have been 10,909 confirmed cases and 256 deaths due to CoVID-19 as of May 11, 2020.[3]

In the field of healthcare, accurate prognosis is essential for efficient management of patients while prioritizing care to the more needy. In order to aid in prognosis, several prediction models have been developed using various methods and tools including machine learning.[4,5] Machine learning is a field of artificial intelligence where computers simulate the processes of human intelligence and can synthesize complex information from huge data sources in a short period of time.[6] Though there have been a few prediction tools on CoVID-19, only a handful have utilized machine learning.[7] To the best of our knowledge, by far there is no publicly available CoVID-19 prognosis prediction model or tool from the general population of confirmed cases using machine learning. We attempt to apply machine learning on the publicly available CoVID-19 data at the community level from South Korea to predict mortality.

Our study had two objectives, (1) predict mortality among confirmed CoVID-19 patients in South Korea using machine learning algorithms, and (2) deploy the best performing algorithm as an open-source online prediction tool for decision-making.

## 2. Material and methods

### 2.1 Patients

Patients for this study were selected from the data shared by Korea Centers for Disease Control and Prevention (KCDC).[3] The timeframe of this study was from the beginning of the detection of the first case (January 20, 2020) through April 30, 2020. In the dataset, there were a total of 3,388 patients. Our inclusion criteria were confirmed CoVID-19 cases with availability of sociodemographic, exposure and diagnosis confirmation features along with the outcome. We excluded patients those had missing features – sex (n=77) and age (n=12), and thus, 3,299 patients were included in the final analysis.

### 2.2 Outcome variable

The outcome variable was mortality and it had a binary distribution – “yes” if the patient died, or “no” otherwise.

### 2.3 Predictors

The predictors were individual patient level socio-demographic and exposure features. They were age group, sex, province, and exposure. There were ten age groups as follows below 10 years, 10-19 years, 20-29 years, 30-39 years, 40-49 years, 50-59 years, 60-69 years, 70-79 years, 80-89 years, 90 years and above. Patients represented all 17 provinces of South Korea (Busan, Chungcheongbuk-do, Chungcheongnam-do, Daegu, Daejeon, Gangwon-do, Gwangju, Gyeonggi-do, Gyeongsangbuk-do, Gyeongsangnam-do, Incheon, Jeju-do, Jeollabuk-do, Jeollanam-do, Sejong, Seoul, and Ulsan). Patients were exposed in several settings, such as nursing home, hospital, religious gathering, call center, community center, shelter and apartment, gym facility, overseas inflow, contact with patients and others.

### 2.4 Statistical Methods

#### 2.4.1 Descriptive Analysis

We performed descriptive analyses of the predictors by respective stratification groups and present the results as numbers and proportions. Potential correlations between predictors were tested with Pearson’s correlation coefficient.

#### 2.4.2 Predictive Analysis

We applied machine learning algorithms to predict mortality among CoVID-19 confirmed cases. Machine learning is a branch of artificial intelligence where computer systems can learn from available data and identify patterns with minimal human intervention.[8] Typically, in machine learning several algorithms are tested on data and performance metrics are used to select the best performing algorithm. We tested five commonly used supervised machine learning algorithms in healthcare research (logistic regression, support vector machine, K neighbor classification, random forest and gradient boosting) to compare algorithm performance efficiency. Logistic regression is best suited for a binary or categorical output. It tries to describe the relationship between the output and predictor variables.[9] In support vector machine (SVM) algorithm, the data is classified into two classes based on the output variable over a hyperplane.[9] The algorithm tries to increase the distance between the hyperplane and the most proximal two data points in each class. SVM uses a set of mathematical functions called kernels. A kernel transforms the inputs to required forms. In our SVM algorithm, we used a linear kernel. K Nearest Neighbors (KNN) is a non-parametric approach that decides the output classification by the majority class among its neighbors.[10] The number of neighbors can be altered to arrive at the best fitting KNN model. For our model, we selected 20 nearest neighbors. Random forest algorithm uses a combination of decision trees.[11] Decision trees are generated by recursively partitioning the predictors. New attributes are sequentially fitted to predict the output. We used an ensemble of 501 decision trees with the trees extended up to a maximum depth of 10.

Gradient boosting (GB) algorithm uses a combination of decision trees.[12] Each decision tree dynamically learns from its precursor and passes on the improved function to the following. Finally, the weighted combination of these trees provides the prediction. A decision tree’s learning from the precursor and the number of subsequent trees can be respectively adjusted using learning rate and number of trees parameters. In our GB model, we used 0.1 learning rate and 51 sequential trees.

#### 2.4.3 Evaluation of the performance of the algorithms

We split the data into training (80 percent) and validation cohorts (20 percent). Initially, the algorithms were trained on the training cohort and then were validated on the validation cohort for determining predictions. The data was passed through a 10-fold cross validation where the data was split into training and validation cohorts at 80/20 ratio randomly ten times. The final prediction came out of the cross-validated estimate. As our data was imbalanced (only 2.1% output were with the condition against 97.9% without), we applied an oversampling technique called synthetic minority oversampling technique (SMOTE) to enhance the learning on the training data.[13,14]

The performance of the algorithms were evaluated for discrimination, calibration and overall performance. Discrimination is the abillity of the algorithm to separate out patients with the mortality risk from those without, where as calibration is the agreement between observed and predicted risk of mortality. An ideal model should have the best of both discrimination and calibration. We tested discriminaiton with area under the receiver operating characteristics curve (AUC) and calibration with accuracy and Matthews correlation coefficient. A receiver operator characteristic (ROC) curve plots the true positive rate on y-axis against the false positive rate on x-axis.[15] AUC is score that measures the area under the ROC curve and it ranges from 0.50 to 1.0 with higher values meaning higher discrimination. Accuracy is a measure of correct classification of death cases as death and survived cases as survived.[15] Matthews correlation coefficient (MCC) is a measure that takes into account all four predictive classes – true positive, true negative, false positive and false negative.[16] It is considered a better measure than accuracy for unbalanced data. Brier score simultaneously account for discrimnation and calibration.[15] A smaller Brier score indicates better performance. In addition, the gradient boosting algorithm was used to estimate the relative contributions of the predictors and draw the variable importance plot.[17]

The statistical analyses were performed using Stata Version 15 (StataCorp LLC. College Station, TX), Python programming language Version 3.7.1 (Python Software Foundation, Wilmington, DE, USA) and R programming language Version 3.6.3 (R Foundation for Statistical Computing, Vienna, Austria). The web application was built using the Shiny package for R and deployed with Shiny server.

## 3. Results

### 3.1 Patient profile

The profile of the patients is presented in Table 1. Out of 3,299 confirmed patients, a slightly more than half were females (56%). Among the age groups, the maximum patients were from 20-29 years (24.3%), followed by 50-59 years (18.1%), 40-49 years (13.8%), 30-39 years (13.3%) and 60-69 years (12.2%). Gyeongsangbuk-do (36.9%), Gyeonggi-do (20.5%) and Seoul (17.1%) provinces together presented the maximum patients. Considering the source/mode of infection, the largest group had unknown mode (40.9%) followed by direct contact with patients (29%) and from overseas (16.8%). According to this available data source, there were 66 deaths accounting for 2.1 percent of the patients.

**Table 1.** Sample characteristics (N=3,299)

| **Variable** | **Number** | **Proportion (%)** |
| --- | --- | --- |
| **Sex** |  |  |
| Female | 1,848 | 56.0 |
| Male | 1,451 | 44.0 |
| **Age group (years)** |  |  |
| Below 10 | 53 | 1.6 |
| 10-19 | 149 | 4.5 |
| 20-29 | 801 | 24.3 |
| 30-39 | 438 | 13.3 |
| 40-49 | 454 | 13.8 |
| 50-59 | 597 | 18.1 |
| 60-69 | 401 | 12.2 |
| 70-79 | 204 | 6.2 |
| 80-89 | 156 | 4.7 |
| 90 and above | 46 | 1.4 |
| **Province** |  |  |
| Busan | 134 | 4.1 |
| Chungcheongbuk-do | 44 | 1.3 |
| Chungcheongnam-do | 143 | 4.3 |
| Daegu | 63 | 1.9 |
| Daejeon | 40 | 1.2 |
| Gangwon-do | 49 | 1.5 |
| Gwangju | 30 | 0.9 |
| Gyeonggi-do | 677 | 20.5 |
| Gyeongsangbuk-do | 1,218 | 36.9 |
| Gyeongsangnam-do | 112 | 3.4 |
| Incheon | 92 | 2.8 |
| Jeju-do | 13 | 0.4 |
| Jeollabuk-do | 17 | 0.5 |
| Jeollanam-do | 15 | 0.5 |
| Sejong | 46 | 1.4 |
| Seoul | 563 | 17.1 |
| Ulsan | 43 | 1.3 |
| **Exposure** |  |  |
| Nursing home | 46 | 1.4 |
| Hospital | 37 | 1.1 |
| Religious gathering | 160 | 4.9 |
| Call center | 112 | 3.4 |
| Community center, shelter and apartment | 50 | 1.5 |
| Gym facility | 34 | 1.0 |
| Overseas inflow | 553 | 16.8 |
| Contact with patients | 957 | 29.0 |
| Others | 1,350 | 40.9 |
| **Outcome** |  |  |
| Survived | 3,230 | 97.9 |
| Died | 69 | 2.1 |

The correlation coefficients among the predictors ranged from -0.12 to 0.03. Using the random forest algorithm, we estimated the relative importance of the predictors (figure 1). Province was the most important predictor followed by age, exposure and sex.

**Figure 1.**
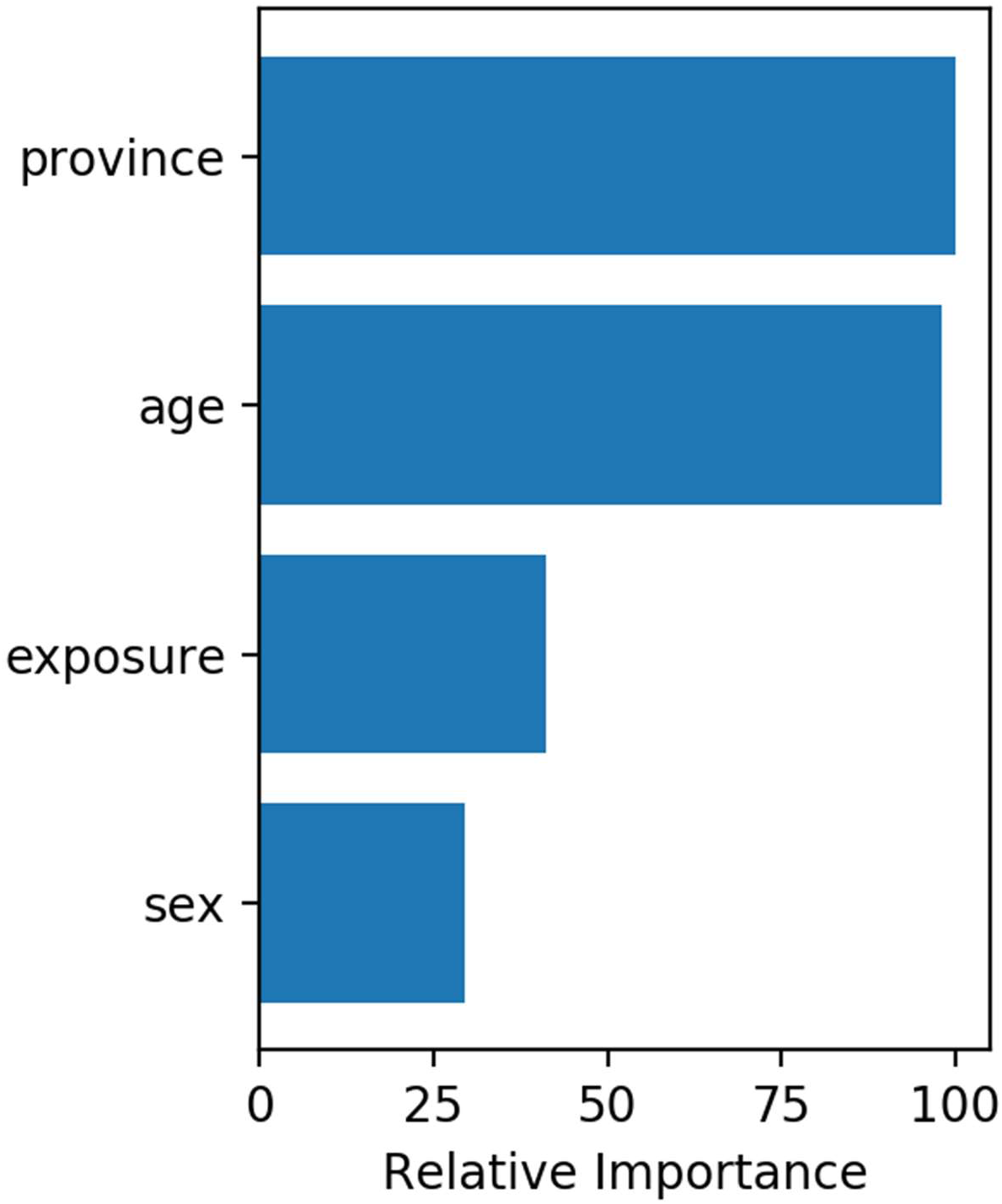
Relative importance of predictors

### 3.2 Performance of the algorithms

Table 2 presents the performance metrics of all algorithms – logistic regression, support vector machine, K nearest neighbor, random forest and gradient boosting. The accuracy of all algorithms was very similar with random forest performing the best (0.981) and logistic regression with the least score (0.971). The area under receiver operating characteristic curve (AUC) ranged from 0.733 to 0.886 with the best score for the random forest algorithm. Similarly, random forest performed the best on Matthews correlation coefficient. It was in the middle for the performance on Brier score. Considering all the performance metrics, random forest was the best performing algorithm.

**Table 2.** Performance of the algorithms with test data.

| **Metrics** | **Logistic**<br>**regression** | **Support**<br>**vector**<br>**machine** | **K nearest**<br>**neighbor** | **Random**<br>**forest** | **Gradient**<br>**boosting** |
| --- | --- | --- | --- | --- | --- |
| Cross-validated<br>accuracy (95% CI) | 0.971<br>(0.954-0.988) | 0.973<br>(0.958-0.988) | 0.979<br>(0.977-0.981) | 0.981<br>(0.972-0.990) | 0.975<br>(0.958-0.992) |
| Area under ROC<br>curve | 0.777 | 0.833 | 0.733 | 0.886 | 0.838 |
| Matthews<br>correlation<br>coefficient | 0.351 | 0.418 | 0.365 | 0.459 | 0.451 |
| Brier score | 0.065 | 0.060 | 0.045 | 0.063 | 0.051 |

### 3.3 Online CoVID-19 mortality risk prediction tool – CoCoMoRP

The best performing model – random forest was deployed as the online mortality risk prediction tool named as “**Co**VID-19 **Co**mmunity **Mo**rtality **R**isk **P**rediction” – CoCoMoRP” (https://ashis-das.shinyapps.io/CoCoMoRP/). Figure 2 presents the user interface of the prediction tool. The web application is optimized to be conveniently used on multiple devices such as desktops, tablets, and smartphones.

**Figure 2.**
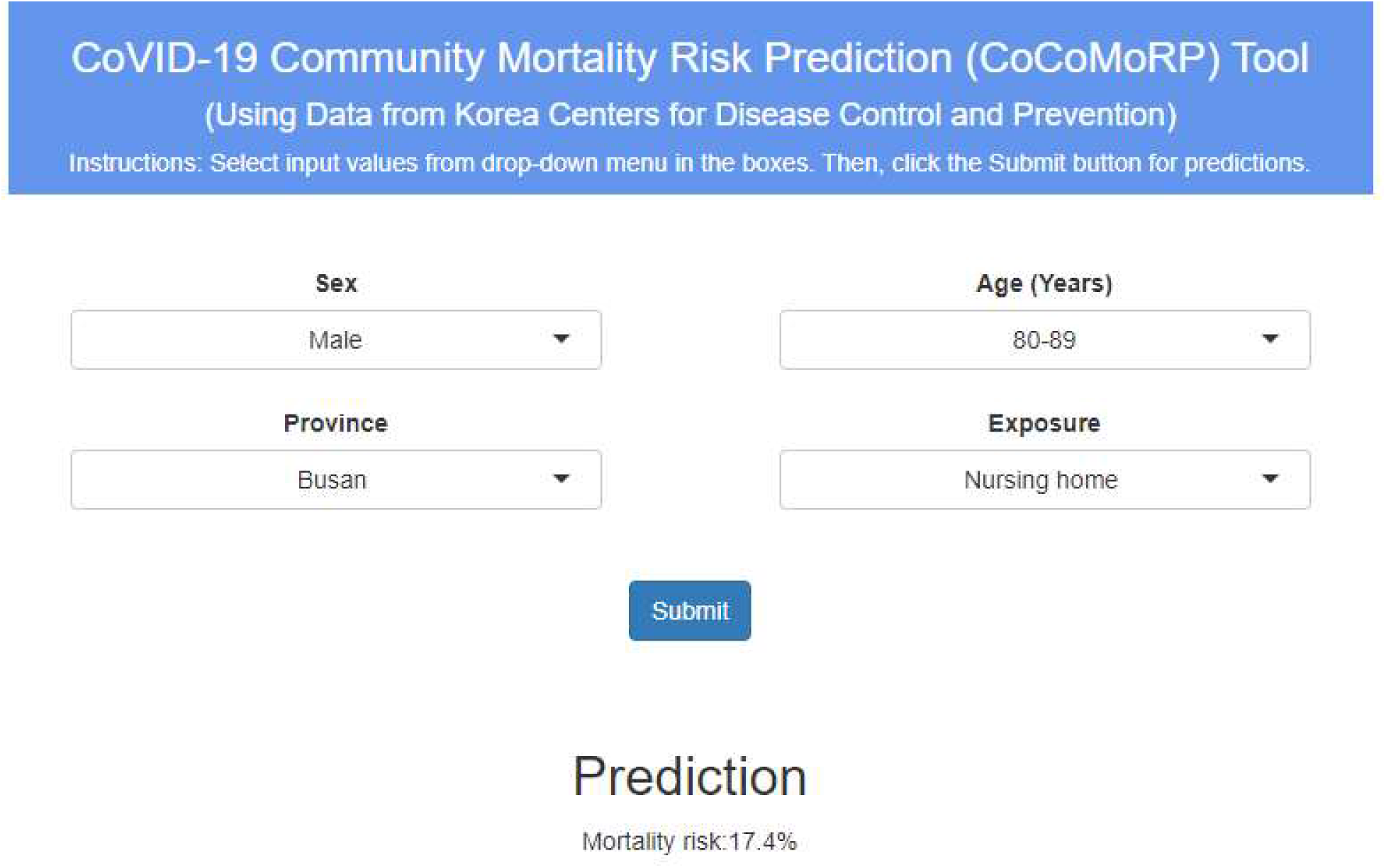
CoCoMORP online **Co**VID-19 **Co**mmunity **Mo**rtality **R**isk **P**rediction tool

The user interface has four boxes to select input features as drop-down menus. The features are sex (two options – male and female), age (ten options – below 10 years, 10-19 years, 20-29 years, 30-39 years, 40-49 years, 50-59 years, 60-69 years, 70-79 years, 80-89 years, 90 years and above), province (all 17 provinces – Busan, Chungcheongbuk-do, Chungcheongnam-do, Daegu, Daejeon, Gangwon-do, Gwangju, Gyeonggi-do, Gyeongsangbuk-do, Gyeongsangnam-do, Incheon, Jeju-do, Jeollabuk-do, Jeollanam-do, Sejong, Seoul, Ulsan), and exposure (nine options – nursing home; hospital; religious gathering; call center; community center, shelter and apartment; gym facility; overseas inflow; contact with patients; and others).

The user has to select one option each from the input feature boxes and click the submit button to estimate the CoVID-19 mortality risk probability in percentages. For instance, the tool gives a CoVID-19 mortality risk prediction of 17.4% for a male patient aged between 80 and 89 years from Busan province with exposure in a nursing home.

## 4. Discussion

The CoVID-19 pandemic is a threat to global health and economic security. Recent evidence for this new disease is still evolving on various clinical and socio-demographic dimensions.[18-20] Simultaneously, health systems across the world are constrained with resources to efficiently deal with this pandemic. We describe the rapid development and deployment of an open-source artificial intelligence informed prognostic tool to predict mortality risk among CoVID-19 confirmed patients using publicly available surveillance data. This tool can be utilized by potential stakeholders such as health providers and policy makers to triage patients at the community level in addition to other approaches.

One major limitation of this tool is unavailability of crucial clinical information on symptoms, risk factors and clinical parameters. Recent research has identified certain symptoms, preexisting illnesses and clinical parameters as strong predictors of prognosis and severity of progression for CoVID-19.[20–22] These crucial pieces of information are not publicly available so far in the surveillance data, so the tool could not be tested to include these features. Inclusion of these additional features may improve the reliability and relevance of the tool. Therefore, we urge the users to balance the predictions from this tool against their own and/or health provider’s clinical expertise and other relevant clinical information.

## 5. Conclusion

We tested multiple machine learning models to accurately predict deaths due to CoVID-19 among confirmed community cases in the Republic of Korea. Using the best performing algorithm, we developed and deployed an online mortality risk prediction tool. To the best of our knowledge, our CoVID-19 community mortality risk prediction tool is the first of its kind. Our tool offers an additional approach to informing decision making for CoVID-19 patients.

## Data Availability

Data are publicly available from Korea CDC

## Authors’ contributions

Conceived and designed this study: Ashis Kumar Das, Shiba Mishra, Saji Saraswathy Gopalan Analyzed and explained the data: Ashis Kumar Das, Shiba Mishra, Saji Saraswathy Gopalan All authors contributed to the writing and approved the final manuscript.

## Acknowledgements

We are grateful to Korea Center for Disease Control and Prevention for making this data publicly available.

## Declaration of competing interest

The authors declare that there is no conflict of interest. The views expressed in the paper are that of the authors and do not reflect that of their affiliations. This particular work was conducted outside of the authors’ organizational affiliations.

## Notes

### Competing Interest Statement

The authors have declared no competing interest.

### Funding Statement

None

